# Innovative Randomized Phase 1 Study and Dosing Regimen Selection to Accelerate and Inform Pivotal COVID-19 Trial of Nirmatrelvir

**DOI:** 10.1101/2022.02.08.22270649

**Authors:** Ravi Shankar P. Singh, Sima S. Toussi, Frances Hackman, Phylinda L. Chan, Rohit Rao, Richard Allen, Lien Van Eyck, Sylvester Pawlak, Eugene P. Kadar, Frances Clark, Haihong Shi, Annaliesa S. Anderson, Michael Binks, Sandeep Menon, Gianluca Nucci, Arthur Bergman

**Affiliations:** Pfizer Worldwide Research, Development and Medical, Cambridge, MA, USA; Pfizer Worldwide Research, Development and Medical, Pearl River, NY, USA; Pfizer Worldwide Research, Development and Medical, Cambridge, UK; Pfizer Global Product Development, Sandwich, UK; Pfizer Clinical Research Unit, Brussels, Belgium; Pfizer Clinical Research Unit, New Haven, CT, USA; Pfizer Worldwide Research, Development and Medical; Groton CT, USA

**Keywords:** COVID-19, SARS-CoV-2, antiviral, nirmatrelvir, PF-07321322, ritonavir

## Abstract

**Background:** COVID-19 is a continued leading cause of hospitalization and death. Safe and efficacious COVID-19 antivirals are needed urgently. Nirmatrelvir (PF-07321332), the first orally bioavailable, SARS-CoV-2 M^pro^ inhibitor against the coronaviridae family, has demonstrated potent preclinical antiviral activity and benign safety profile.

**Methods:** We report safety, tolerability, and pharmacokinetic data of nirmatrelvir with and without ritonavir as a pharmacokinetic enhancer, from an accelerated randomized, double-blind, placebo-controlled, phase 1 study. Two interleaving single-ascending dose (SAD) cohorts were evaluated in a 3-period crossover. Multiple-ascending dose (MAD) with nirmatrelvir/ritonavir twice daily (BID) dosing was evaluated over 10 days in 5 parallel cohorts. Safety was assessed, including in a supratherapeutic exposure cohort. Dose and dosing regimen for clinical efficacy evaluation in phase 2/3 clinical trials were supported by integrating modelling and simulations of SAD/MAD data with nonclinical data and a quantitative systems pharmacology model (QSP).

**Results:** In SAD, MAD, and supratherapeutic exposure cohorts, nirmatrelvir/ritonavir was safe and well tolerated. Nirmatrelvir exposure and half-life were considerably increased by ritonavir, enabling selection of nirmatrelvir/ritonavir dose and regimen for phase 2/3 trials (300/100 mg BID), to achieve concentrations continuously above those required for 90% inhibition of viral replication in vitro. The QSP model suggested that a 5-day regimen would significantly decrease viral load in SARS-CoV-2-infected patients and prevent development of severe disease, hospitalization, and death.

**Conclusions:** An innovative and seamless trial design expedited establishment of phase 1 safety and pharmacokinetics of nirmatrelvir/ritonavir, enabling high confidence in phase 2/3 dose selection and accelerated pivotal trials’ initiation. NCT04756531

## INTRODUCTION

COVID-19 is a continued threat to public health worldwide more than 2 years after its emergence. Despite availability of effective vaccines,^1, 2^ infection rates remain high, and COVID-19 continues to be a leading cause of hospitalization and death.^3^ Intravenously administered monoclonal antibodies were the first outpatient therapies to be made available under emergency use authorization (EUA) to patients at high risk of severe COVID-19.^4–7^ However, logistical constraints could affect administration, and reduced effectiveness has been observed for some monoclonal antibodies, leading the US Government to recommend not using them for some SARS-CoV-2 variants.^4, 8^ At the time of this study, no therapies were available to treat COVID-19 in community settings; an urgent need therefore exists for safe and efficacious orally administered therapies, particularly for individuals at high risk of severe illness.

The coronaviridae family, including SARS-CoV-2, encodes 2 proteases from which functional proteins are generated through proteolysis, one of which is the main protease (M^pro^).^9,10^ M^pro^ is a promising target for viral inhibitors because it is crucial for processing viral polyproteins into functional units,^11^ is highly conserved across SARS-CoV-2 and other coronaviruses,^12, 13^ and has limited potential for off-target activity with no identified human analogs.^14^

Nirmatrelvir (PF-07321332), designed to be orally administered, demonstrated potent and specific inhibition of M^pro^ enzyme activity and anti-viral activity across a diverse spectrum of coronaviruses in preclinical assays.^15^ Oral administration in a murine SARS-CoV-2 model demonstrated dose-dependent reduction of pulmonary viral titers and reduced tissue pathology.^15^ At the time of study initiation, it was not clear if nirmatrelivir concentrations several fold over the in vitro 90% effective concentration (EC_90_) could be maintained in humans; however since nirmatrelvir is primarily metabolized by cytochrome P450 3A4 (CYP3A4), dosing with a pharmacokinetic enhancing agent was included in the protocol to ensure that efficacious concentrations could be achieved.^15^

In response to the public health crisis resulting in an urgent need for COVID-19 therapeutics, we used an innovative and seamless operational approach incorporating a flexibly written protocol that allowed study conduct to adapt to emerging data within the study without the need for a protocol amendment. We simultaneously utilized model informed drug development (MIDD) to expedite dose selection and inform the design of phase 2/3 clinical studies. This study in healthy adults examined safety, tolerability, and pharmacokinetics of nirmatrelvir, with and without ritonavir as a pharmacokinetic enhancer to prevent rapid metabolism of nirmatrelvir by CYP3A4, thus ensuring that free concentrations well above in vitro EC_90_ are maintained in >90% of the population to achieve the optimal therapeutic effect.

## METHODS

### Objectives, Participants, Oversight

This 5-part study (NCT04756531) assessed nirmatrelvir safety, tolerability, and pharmacokinetics following single-ascending doses (SAD) including food effect, multiple-ascending doses (MAD), and at supratherapeutic exposure in healthy 18‒60-year-olds. MAD evaluation also included a Japanese cohort. Relative bioavailability and food effect of an oral tablet formulation without pharmacokinetic enhancer, and nirmatrelvir metabolism and excretion, were evaluated in 2 additional open-label cohorts (to be reported separately). SAD, MAD, and supratherapeutic exposure cohorts were randomized and double-blinded (to participants and investigators; open to a subset of the blinded study team for efficient decision-making). Key inclusion/exclusion criteria, ethical study conduct, study responsibilities, operational conduct, and methods to ensure blinding are summarized in the **Supplementary Information**.

### Randomization and Study Treatment

Within 2 cohorts in SAD, participants were randomly assigned using a randomization schedule to nirmatrelvir or placebo in an interleaving 3-period crossover, where dose escalations were alternated between the 2 cohorts (**Figure 1A**). In each period, participants received nirmatrelvir suspension or placebo, with ≥5 day washout between doses. In the first 3 dosing periods, nirmatrelvir administered fasted without ritonavir was escalated from 150 mg to 1500 mg. In the next 2 dosing periods, nirmatrelvir administered fasted was escalated from 250 mg to 750 mg while coadministered with ritonavir 100 mg. The final dosing period evaluated food effect on pharmacokinetics of nirmatrelvir 250 mg with ritonavir 100 mg. Where applicable, ritonavir was administered at −12, 0, and 12 hours relative to nirmatrelvir. For each dose escalation decision, preliminary cumulative safety, tolerability, and pharmacokinetic data from previous dosing periods were evaluated.

**Figure 1.**
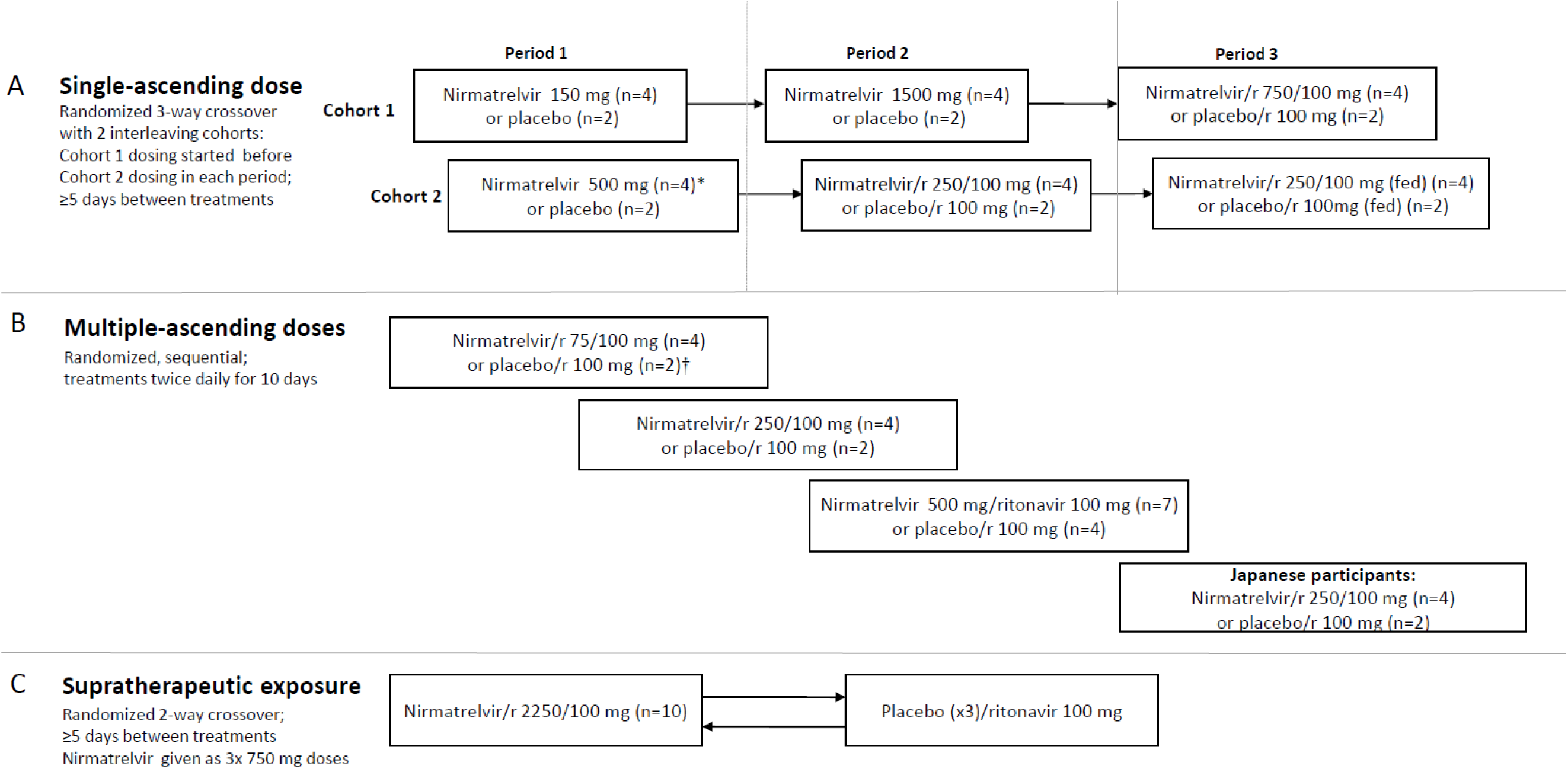
Study design and numbers of participants treated. *One participant in the nirmatrelvir 500 mg group during the single-ascending dose part withdrew due to an adverse event. †One participant in the placebo/ritonavir group during the multiple-ascending dose part discontinued due to participant withdrawal. In this study, which was conducted at the Pfizer Clinical Research Unit, nirmatrelvir was given as an oral suspension. Unless otherwise stated, nirmatrelvir was given under fasting conditions. During the supratherapeutic exposure assessment, participants remained in the clinical trials unit during the washout period. r=ritonavir.

In MAD, participants were randomly assigned to escalating nirmatrelvir dose levels (75, 250, 500 mg) or placebo, all administered twice daily (BID) as a suspension for 10 days with ritonavir 100 mg, under fasting conditions (**Figure 1B**). For each dose escalation decision, preliminary cumulative safety, tolerability, and pharmacokinetic data from previous treatment cohorts were evaluated. After dose escalation, a cohort of Japanese participants was randomized to receive nirmatrelvir 250 mg or placebo, with ritonavir 100 mg, under fasting conditions BID for 10 days.

To assess supratherapeutic exposure in the presence of solubility-limited absorption, split dosing was used, where participants were randomly assigned to receive 2250 mg of nirmatrelvir (administered as 3×750 mg doses at 0, 2 and 4 hours) or matching placebo in a 2-way crossover with ≥5 days washout between treatments (**Figure 1C**). Ritonavir was administered at −12, 0, and 12 hours relative to the first nirmatrelvir or placebo dose.

The **Supplementary Information** provides additional details regarding randomization, including the method used to generate the random allocation sequence and the type of randomization.

### Safety and Tolerability

Safety and tolerability evaluations included adverse event (AE) and serious AE (SAE) assessments between signing of informed consent until 28 days after study treatment administration. AEs are presented by the MedDRA v24.0 term. Vital signs, electrocardiograms (ECGs), and laboratory tests were monitored at prespecified time points.

### Bioanalytical Methods

Human plasma samples were analyzed utilizing a validated LC-MS/MS assay. The assay consisted of protein precipitation in conjunction with reversed phase liquid chromatography coupled to positive ion electrospray tandem quadrupole mass spectrometry. Precipitation was conducted by aliquoting 100 µL of plasma sample to a 96-well propylene plate then adding acetonitrile or working internal standard in acetonitrile (300 µL). The plate was mixed and centrifuged at 3000 rpm. Supernatant (50.0 µL) from each well was transferred to a new 96-well propylene plate and diluted with 500 µL of 60:40 (v:v) water:acetonitrile containing 0.1% formic acid, mixed and injected on the LC-MS/MS system. The analytical column employed was a Waters Acquity UPLC BEH C18, 1.7 µm, 2.1 × 50 mm operated at ambient temperature. Mobile phases A and B consisted of water containing 0.1% formic acid and acetonitrile containing 0.1% formic acid, respectively, with a flow rate of 0.600 mL/min. The gradient program was initiated with a (70:30) mobile phase A:B and held for 0.1 minutes, followed by a linear gradient to (60:40) mobile phase A:B to 5.0 minutes. A column wash was applied at (15:85) mobile phase A:B to 6.0 minutes, followed by re-equilibration at (70:30) mobile phase A:B to 7.0 minutes. A Sciex API5500 tandem quadrupole mass spectrometer equipped with a Turbo V Ion Source was operated in electrospray (TurbolonSpray^®^) positive mode. The parent to product ion transitions 500.5/110.3 and 509.5/109.9 were utilized for PF-07321332 and PF 07818226 (PF-07321332 internal standard), respectively. Peak areas of the analytes and internal standards were determined by Analyst data processing software (version 1.7.2). These responses were imported into Watson LIMS system (version 7.6.1) and a calibration curve was constructed using peak area ratios of the calibration samples and applying weighted (1/X^2^) linear least squares regression analysis. The method was validated over the range of 10.0‒50,000 ng/mL.

### Pharmacokinetic Evaluations

Nirmatrelvir plasma pharmacokinetic parameters as described in **Table S1** following single and multiple oral doses, and urine pharmacokinetics following multiple oral doses were derived from concentration-time data using standard noncompartmental methods.

### Dose and Duration Selection for Phase 2/3

Consistent with the literature for other protease inhibitors,^16, 17^ robust antiviral efficacy is predicted when unbound plasma concentrations are sustained above multiples of the EC_90_ (ie, concentration at which 90% inhibition of viral replication occurs) for the entire dosing interval. The unbound target minimum plasma concentration (C_min_) to be maintained corresponded to the in vitro drug concentration at which 90% inhibition of SARS-CoV-2 viral replication is observed (adjusting for plasma protein binding the total plasma EC_90_=292 ng/mL).^15^ A population pharmacokinetic model of nirmatrelvir with ritonavir 100 mg was built during the dose escalation based on the preliminary SAD/MAD data from healthy adults in this study to project distribution of expected nirmatrelvir concentrations at different doses and regimens. Consistent with the anticipated phase 2/3 dosing regimen, only the pharmacokinetic data collected from the nirmatrelvir/ritonavir treatment arms were included in the population pharmacokinetic model. This enabled selection of a phase 2/3 dose that would result in C_min_ above EC_90_ in the vast majority (≥90%) of future trial participants.

In the population pharmacokinetic model, logarithmically transformed plasma nirmatrelvir concentration versus time data were analyzed with NONMEM, version 7.5.0 and the FOCE method with interaction. A 1- or 2-compartment model with first-order elimination and first-order absorption were tested as the structural model. The base model included an allometric model of baseline body weight on apparent clearances (CL/F and Q/F) and volumes (V2 and V3) with exponentials fixed to 0.75 and 1, respectively. Inter-individual variability and inter-occasion variability in the pharmacokinetic parameters were assumed to be log normally distributed and modelled using multiplicative exponential random effects. Models with and without covariance for random effects were tested. Residual random effects were described with a combined proportional and additive model in the log domain. Food effect on k_a_ and relative bioavailability (F1) and time-dependent change in clearance were evaluated.

Simulations were performed utilizing the preliminary population pharmacokinetic model with nirmatrelvir doses of 100‒500 mg by 100 mg increments with ritonavir given BID for 5 days (single dose on Day 5) assuming no missing doses. Interindividual variability in nirmatrelvir clearance was assumed to be 60% to account for expected increases in interindividual variability in phase 2/3 studies relative to this phase 1 study. Simulated nirmatrelvir plasma concentration profiles were used to calculate the percentage of participants achieving a trough concentration (C_min_) of at least the in vitro EC_90_.

To select the treatment duration, a quantitative systems pharmacology (QSP) model capable of describing viral dynamics over time was used to predict potential viral load reduction as a measure of efficacy.^18, 19^ The model was updated to include expected nirmatrelvir pharmacokinetic data at the proposed phase 2/3 dose, preclinical data from a mouse model of SARS-CoV-2, and publicly available viral load data from randomized controlled trials.^20–22^ Specifically, to predict the efficacy and optimal dosing regimen of nirmatrelvir, the QSP model was updated to incorporate: (1) the mean simulated pharmacokinetic profile of nirmatrelvir/ritonavir 300 mg/100 mg BID 5 day and 10 day regimens from the population pharmacokinetic model described in the preceding section; (2) preclinical data on nirmatrelvir pharmacology in a mouse model of SARS-CoV-2 that was used to estimate the in vivo potency of nirmatrelvir with the QSP model; and (3) a virtual population (N=502) that matched the placebo and treatment response of viral load and severity as reported in publicly available data.^20–23^

A virtual population was simulated to predict the effects of different dosing durations (ie, 5 or 10 days of dosing),^24^ with a symptomatic outpatient COVID-19 population and dosing 4 days post viral load peak/symptom onset assumed.^20^ The model was used to predict the influence of the 5 day and 10 day dosing regimens on the viral load time course at the estimated in vivo potency of nirmatrelvir. Moreover, given the uncertainty on the clinical potency of nirmatrelvir prior to phase 2/3 dose and regimen selection, sensitivity analysis was conducted to evaluate the impact of a range of potencies simulated in the model (**Figure S1**). For this sensitivity analysis, the viral load lowering efficacy was predicted at the start of Day 7 and Day 10 post treatment.

### Statistical Analysis

The study did not include statistical hypotheses. Safety endpoints were summarized in the safety population (**Table S2**) as counts and percentages. SAD and MAD were to include 6 participants per cohort (nirmatrelvir, 4; placebo, 2). For supratherapeutic exposures, a sample size of 12 participants (with ≥10 completers) was chosen.

Pharmacokinetic parameters were assessed in the pharmacokinetic parameter population (**Table S2**). No formal inferential statistics were applied to pharmacokinetic data apart from comparisons of food effect in SAD (**Supplementary Information**).

## RESULTS

### Participants

Dosing for this study began on March 2, 2021 and dose selection was completed on April 30, 2021. The study randomized 13, 29, and 10 healthy adults in the SAD, MAD, and supratherapeutic parts of the study, respectively. One participant discontinued during SAD due to an AE; 1 participant discontinued during MAD due to participant withdrawal. All 10 participants assessed for supratherapeutic exposure completed the study (**Figure 1**). Demographic characteristics are summarized in **Table 1**.

**Table 1.**
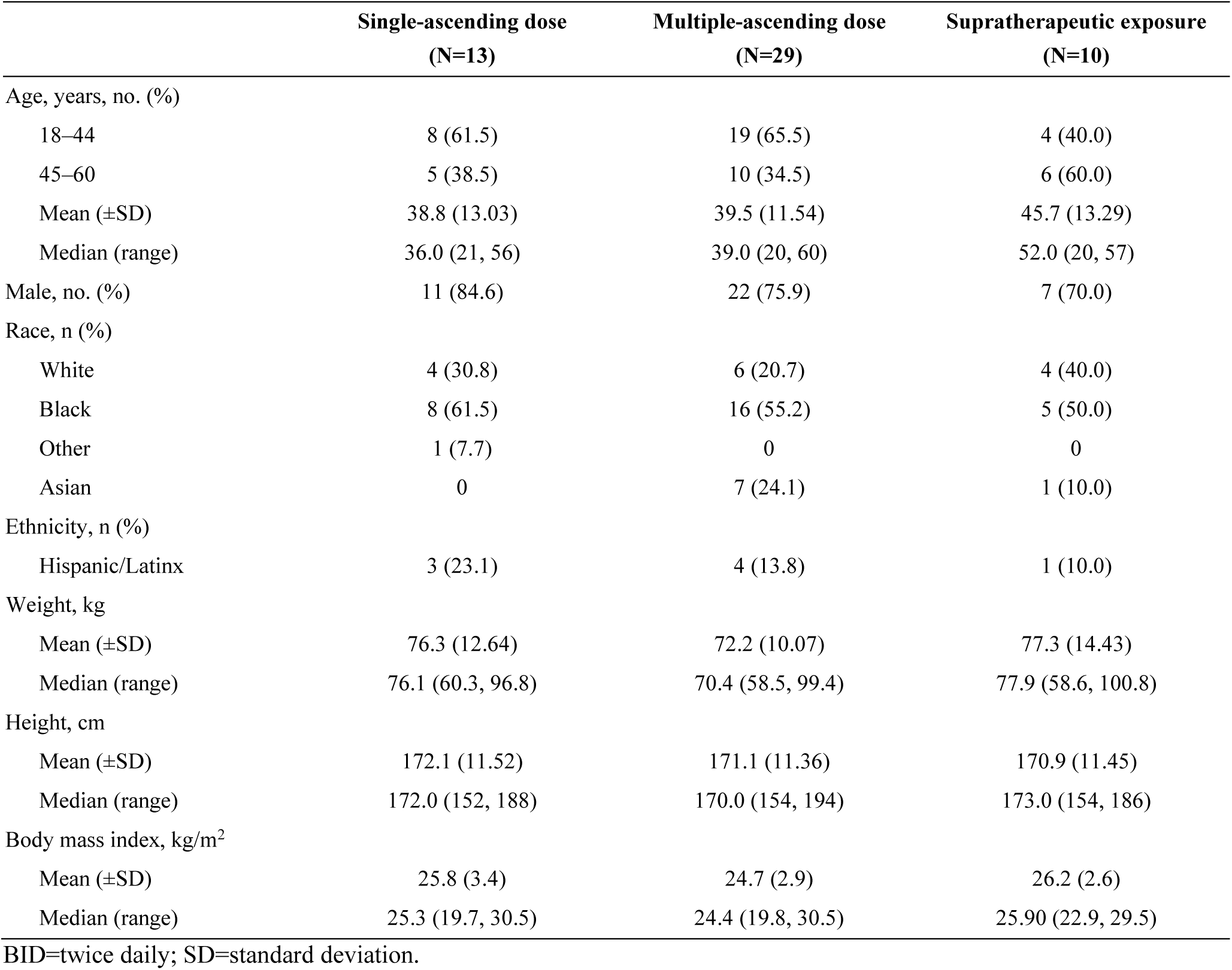
Demographic and clinical characteristics.

### Safety

No participant had an SAE, severe AE, or the dose reduced or temporarily discontinued because of AEs. One participant in SAD discontinued from the study because of a positive COVID-19 test. This was a protocol-specified test occurring on Day 4 and the participant was asymptomatic. All participants in MAD completed 10 days of treatment apart from 1 participant who withdrew after receiving placebo/ritonavir 100 mg BID (fasted) for 7 days.

In SAD, no AEs were considered treatment-related **(Table S3),** and all AEs were mild. Five treatment-related events of blood thyroid stimulating hormone (TSH) increased occurred in MAD, and 2 of these events occurred in the placebo groups **(Table S4)**. Three treatment-related events of dysgeusia occurred, and all were in the treated groups. All Japanese participants in MAD reported AEs and 3 were considered treatment-related (2 participants with mild blood TSH increased, including 1 on placebo; 1 participant with mild dysgeusia). At supratherapeutic exposure, 1 participant each had 1 treatment-related AE: nausea with treatment and vomiting with placebo. Overall, no safety concerns were identified with administration of nirmatrelvir, with or without ritonavir. There was no evidence of a dose-related increase in AEs. No clinically meaningful changes in laboratory values (≥ grade 2), vital sign measurements, or ECG measurements were observed.

### Pharmacokinetics

Pharmacokinetic parameters for nirmatrelvir calculated in SAD are summarized in **Figure 2A, Table 3**, and **Table S5**. Following single-dose administration of nirmatrelvir as an oral suspension at doses of 150 mg, 500 mg, and 1500 mg without ritonavir under fasted conditions, less than dose proportional increases in nirmatrelvir exposure were observed. Ritonavir considerably increased nirmatrelvir exposure. Following administration of nirmatrelvir 250 mg or 750 mg enhanced with 100 mg ritonavir, a dose-related increase in nirmatrelvir was observed albeit in a less than dose-proportional manner. Following administration of a 250 mg oral suspension of nirmatrelvir with ritonavir 100 mg under fed and fasted conditions, no meaningful change in exposure was observed (∼1.5% increase in AUC and ∼15% increase in C_max_).

**Figure 2.**
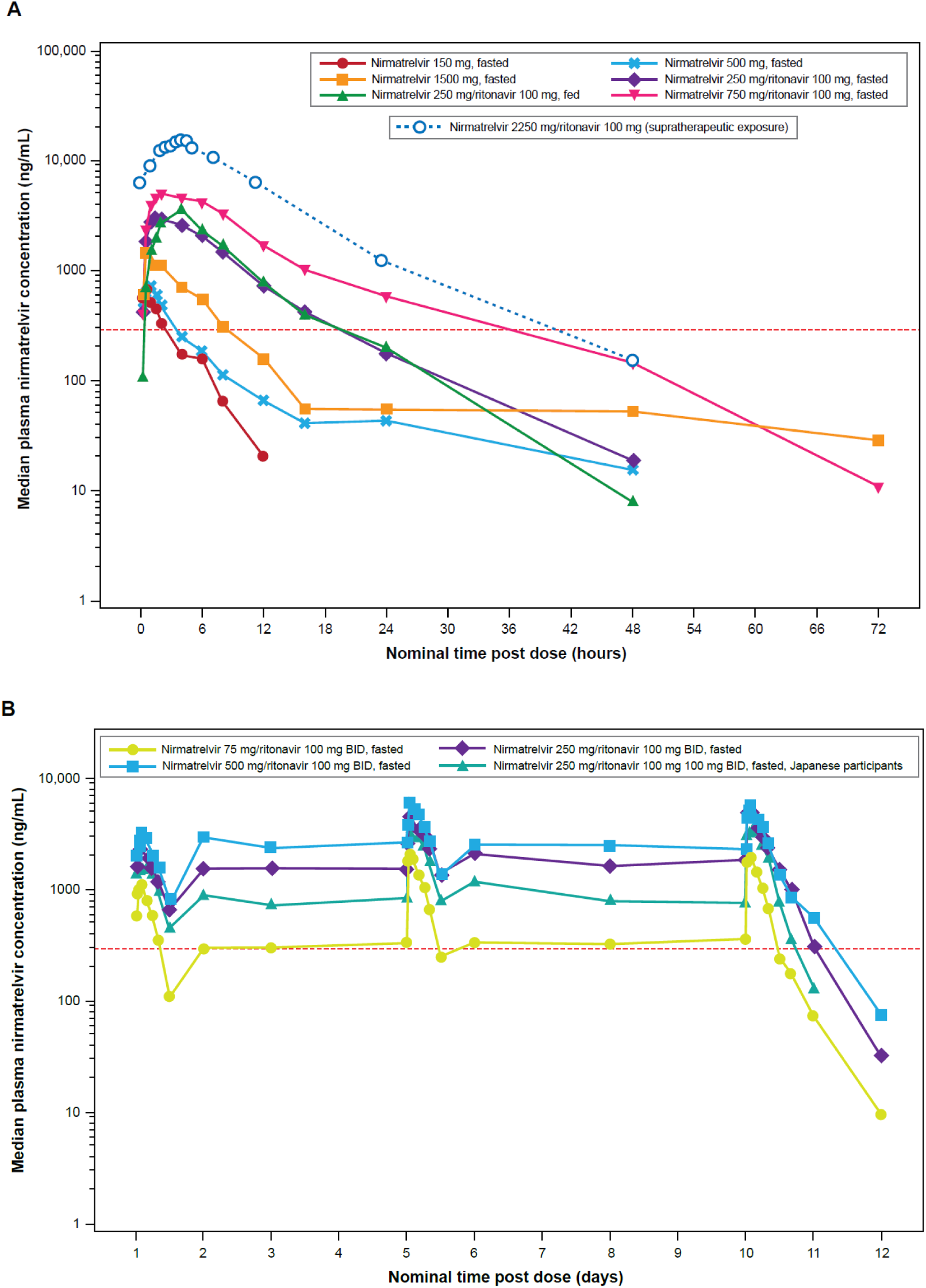
Median plasma nirmatrelvir concentration-time profiles (semi-log scales) for single-ascending dose and supratherapeutic exposure cohorts (A) and multiple-ascending dose cohort (B). For summary statistics, values below the lower limit of quantification (10 ng/mL) were set to zero. In the supratherapeutic exposure assessment, nirmatrelvir was administered as 3×750 mg doses at 0, 2, and 4 hours. In the single-ascending dose assessments where applicable and supratherapeutic exposure assessments, ritonavir was dosed at 100 mg at –12 hours, 0 hours, and 12 hours after dosing. In the multiple-ascending dose assessment, ritonavir was dosed at 100 mg twice daily. The red dotted line is EC_90_ of 292 ng/mL. BID=twice daily; EC_90_=concentration at which 90% inhibition of viral replication is observed.

**Table 2.** Single-ascending dose assessment of plasma nirmatrelvir pharmacokinetic parameters: descriptive summary (A) and statistical summary of food effect (B)

| **A. Descriptive summary** |  |  |  |  |  |  |
| --- | --- | --- | --- | --- | --- | --- |
| **Parameter (unit)\*** | **Nirmatrelvir 150 mg**<br>**Fasted (N=4)** | **Nirmatrelvir 500 mg**<br>**Fasted (N=4)** | **Nirmatrelvir 1500 mg**<br>**Fasted (N=4)** | **Nirmatrelvir/r**<br>**250/100 mg**<br>**Fasted (N=4)** | **Nirmatrelvir/r**<br>**250/100 mg**<br>**Fed (N=4)** | **Nirmatrelvir/r**<br>**750/100 mg**<br>**Fasted (N=4)** |
| N1, N2* | 4, 3 | 4, 2 | 4, 0 | 4, 4 | 4, 4 | 4, 4 |
| AUC<sub>inf</sub>, ng·h/mL | 2247 (42) | 5480, 5450† | NR | 28,220 (14) | 28,640 (17) | 66,760 (45) |
| C<sub>max</sub>, ng/mL | 667.7 (28) | 674.4 (38) | 1538 (32) | 2882 (25) | 3323 (13) | 5086 (25) |
| t<sub>1/2</sub>, h | 2.023 ± 0.54556 | 18.5, 25.6† | NR | 6.935 ± 1.0794 | 6.005 ± 1.6502 | 12.86 ± 8.4196 |
| T<sub>max</sub>, h | 0.634 (0.550–1.50) | 1.00 (0.517–1.00) | 1.00 (0.533–2.00) | 2.75 (1.50–4.00) | 4.00 (4.00–4.00) | 2.00 (1.50–4.00) |
| **B. Statistical summary – food effect** |  |  |  |  |  |  |
| **Parameter (unit)\*** | **Nirmatrelvir/r**<br>**250/100 mg fed (test)** | **Nirmatrelvir/r**<br>**250/100 mg fasted (test)** | **Ratio (%) (test/reference)**<br>**of adjusted geometric means\*** |  | **90% CI (%) for ratio‡** |  |
| AUC<sub>inf</sub>, ng·h/mL | 28,640 | 28,220 | 101.5 |  | (89.57, 115.07) |  |
| C<sub>max</sub>, ng/mL | 3323 | 2882 | 115.3 |  | (99.36, 133.79) |  |

The statistical summary of food effect used mixed effect models with treatment as a fixed effect and participant as a random effect, which were applied to the natural log transformation of AUC<sub>inf</sub> and C<sub>max</sub> separately using participants with data from both periods only.
\* N1 is the number of participants contributing to the summary statistics and N2 is the number of participants where t<sub>1/2</sub> and AUC<sub>inf</sub> were determined.
† Where fewer than 3 participants had evaluable measurements, individual values are listed.
‡ Ratios (and 90% CIs) are expressed as percentages.
The parameters are defined in **Table S1**. Additional results are in **Table S5**.
CI=confidence interval; NR=not reported; r=ritonavir.

**Table 3.**
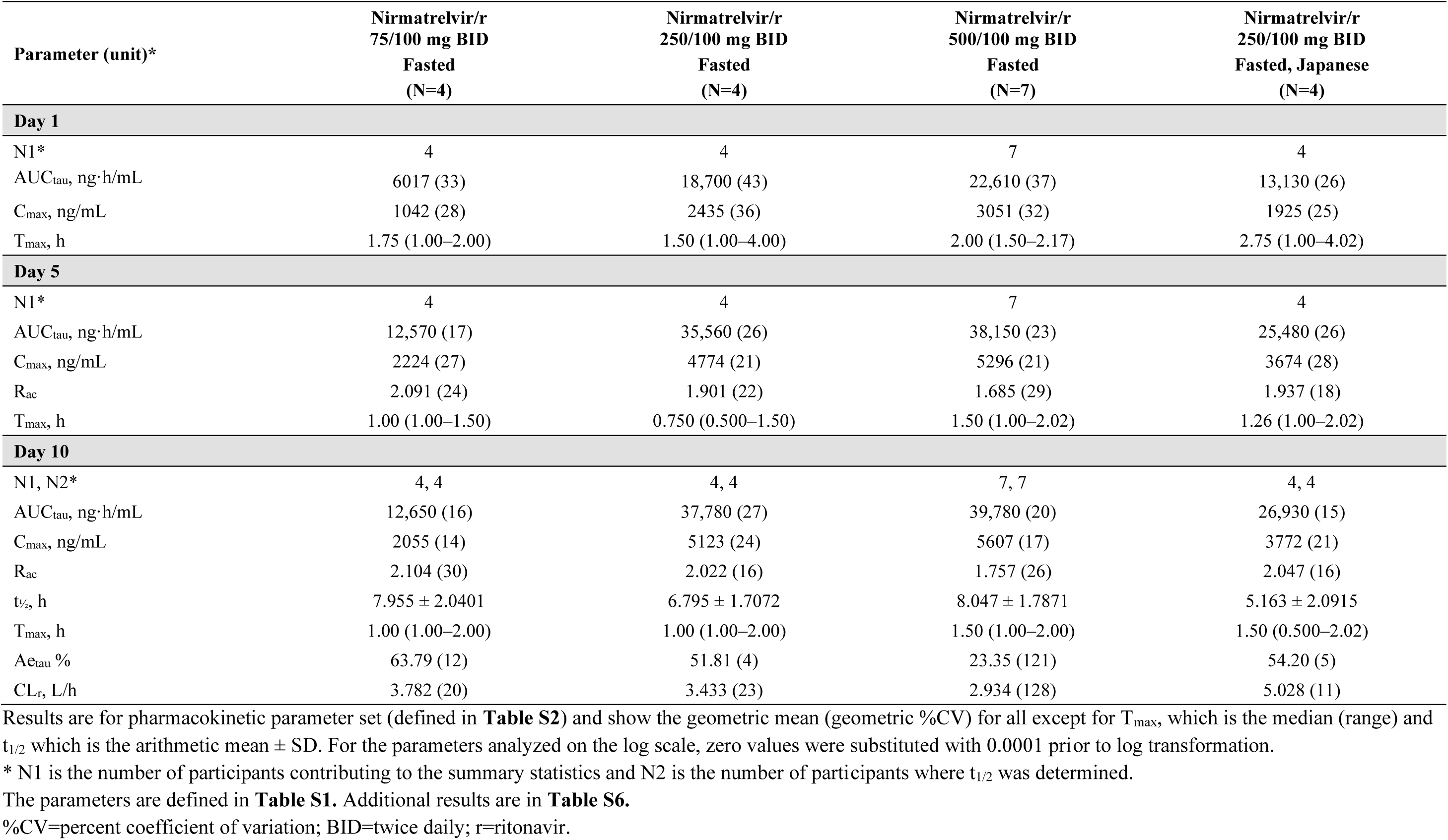
Multiple-ascending dose assessment: descriptive summary of plasma and urine nirmatrelvir pharmacokinetic parameters.

| Parameter (unit)* | Nirmatrelvir/r<br>75/100 mg BID<br>Fasted<br>(N=4) | Nirmatrelvir/r<br>250/100 mg BID<br>Fasted<br>(N=4) | Nirmatrelvir/r<br>500/100 mg BID<br>Fasted<br>(N=7) | Nirmatrelvir/r<br>250/100 mg BID<br>Fasted, Japanese<br>(N=4) |
| --- | --- | --- | --- | --- |
| **Day 1** |  |  |  |  |
| N1* | 4 | 4 | 7 | 4 |
| AUC<sub>tau</sub>, ng·h/mL | 6017 (33) | 18,700 (43) | 22,610 (37) | 13,130 (26) |
| C<sub>max</sub>, ng/mL | 1042 (28) | 2435 (36) | 3051 (32) | 1925 (25) |
| T<sub>max</sub>, h | 1.75 (1.00–2.00) | 1.50 (1.00–4.00) | 2.00 (1.50–2.17) | 2.75 (1.00–4.02) |
| **Day 5** |  |  |  |  |
| N1* | 4 | 4 | 7 | 4 |
| AUC<sub>tau</sub>, ng·h/mL | 12,570 (17) | 35,560 (26) | 38,150 (23) | 25,480 (26) |
| C<sub>max</sub>, ng/mL | 2224 (27) | 4774 (21) | 5296 (21) | 3674 (28) |
| R<sub>ac</sub> | 2.091 (24) | 1.901 (22) | 1.685 (29) | 1.937 (18) |
| T<sub>max</sub>, h | 1.00 (1.00–1.50) | 0.750 (0.500–1.50) | 1.50 (1.00–2.02) | 1.26 (1.00–2.02) |
| **Day 10** |  |  |  |  |
| N1, N2* | 4, 4 | 4, 4 | 7, 7 | 4, 4 |
| AUC<sub>tau</sub>, ng·h/mL | 12,650 (16) | 37,780 (27) | 39,780 (20) | 26,930 (15) |
| C<sub>max</sub>, ng/mL | 2055 (14) | 5123 (24) | 5607 (17) | 3772 (21) |
| R<sub>ac</sub> | 2.104 (30) | 2.022 (16) | 1.757 (26) | 2.047 (16) |
| t<sub>1/2</sub>, h | 7.955 ± 2.0401 | 6.795 ± 1.7072 | 8.047 ± 1.7871 | 5.163 ± 2.0915 |
| T<sub>max</sub>, h | 1.00 (1.00–2.00) | 1.00 (1.00–2.00) | 1.50 (1.00–2.00) | 1.50 (0.500–2.02) |
| Ae<sub>tau</sub> % | 63.79 (12) | 51.81 (4) | 23.35 (121) | 54.20 (5) |
| CL<sub>r</sub>, L/h | 3.782 (20) | 3.433 (23) | 2.934 (128) | 5.028 (11) |

\* N1 is the number of participants contributing to the summary statistics and N2 is the number of participants where t<sub>1/2</sub> was determined.
The parameters are defined in **Table S1**. Additional results are in **Table S6**.
%CV=percent coefficient of variation; BID=twice daily; r=ritonavir.

Pharmacokinetic parameters for nirmatrelvir calculated in MAD are summarized in **Figure 2B, Table 4**, and **Table S6**. Following multiple-dose administration of nirmatrelvir/ritonavir at 75/100 mg, 250/100 mg, and 500/100 mg BID doses under fasted conditions, nirmatrelvir exposure on Days 1, 5, and 10 increased in a less than dose-proportional manner across the doses studied. Plasma nirmatrelvir levels increased approximately 2-fold after the first dose, with steady-state concentrations achieved by Day 2 for all doses and treatments and maintained on Days 5 and 10. Urinary recovery of unchanged nirmatrelvir was substantial, indicating kidney as a major organ in nirmatrelvir elimination in the presence of ritonavir. Nirmatrelvir renal clearance was similar across all doses. Nirmatrelvir exposure, accumulation, and urinary recovery were not meaningfully different in Japanese participants across all days.

**Table 4.** Descriptive summary of plasma nirmatrelvir pharmacokinetic parameters for supratherapeutic effect.

| Parameter (unit) | Nirmatrelvir/r 2250*/100 mg<br>(N=10) |
| --- | --- |
| N1, N2† | 10, 10 |
| AUC<sub>inf</sub>, ng·h/mL | 188,800 (35) |
| C<sub>max</sub>, ng/mL | 15,940 (27) |
| T<sub>max</sub>, h | 5.00 (3.02–6.03) |

\* Given as 3 doses of 750 mg at 0, 2, and 4 hours.
† N1 is the number of participants contributing to the summary statistics and N2 is the number of participants where AUC<sub>inf</sub> was determined.
The parameters are defined in **Table S1**. Additional results are in **Table S7**.

Pharmacokinetic parameters for nirmatrelvir calculated in the supratherapeutic assessment are summarized in **Figure 2A**, **Table 5**, and **Table S7**. Following a supratherapeutic oral dose of nirmatrelvir as a 2250-mg suspension (dosed as 3 split doses of 750 mg administered at 0, 2 and 4 hours) enhanced with 100 mg ritonavir, the mean C_max_ was 5 times higher than the mean C_max_ observed in the nirmatrelvir/ritonavir 500/100 mg BID cohort on Day 1 in the MAD part of the study.

### Dose and Duration Selection in Phase 2/3

The pharmacokinetics of nirmatrelvir with ritonavir 100 mg following oral administration was adequately characterized by a 2-compartment disposition model with first-order absorption. Separate power functions were used to describe the dose-effect on k_a_ and F1. The parameter estimates for a nirmatrelvir dose of 300 mg with ritonavir were clearance 8.2 L/h, volume of distribution 111 L, and k_a_ 1.11 h^-1^. No obvious time-dependent change in nirmatrelvir clearance was noted. A high-fat meal reduced k_a_ by approximately 50% but had minimal effect on F1 (reduced by 6.5%). Considering its impact on C_min_ would be minimal, and the inclusion of inter-individual and inter-occasion variability on k_a_, food effect was not included in the final model for subsequent simulation.

Concentrations were simulated for a sample size commensurate with phase 2/3 studies for a range of nirmatrelvir doses. With the population pharmacokinetic model based on the preliminary data from this clinical study, the nirmatrelvir/ritonavir 300/100 mg BID simulation showed that >90% of future trial participants would achieve the target C_min_ above EC_90_ after the first and subsequent dose and with interindividual variability in clearance inflated to 60% (**Table S8**). The projected median C_min_ on Day 1 and at steady state was 987 ng/mL and 1800 ng/mL, respectively, which are approximately 3 and 6 times higher than the in vitro EC_90_, and therefore projected to achieve efficacy. Simulated steady-state (Day 5) C_min_ at different doses is shown in **Figure 3A**.

**Figure 3.**
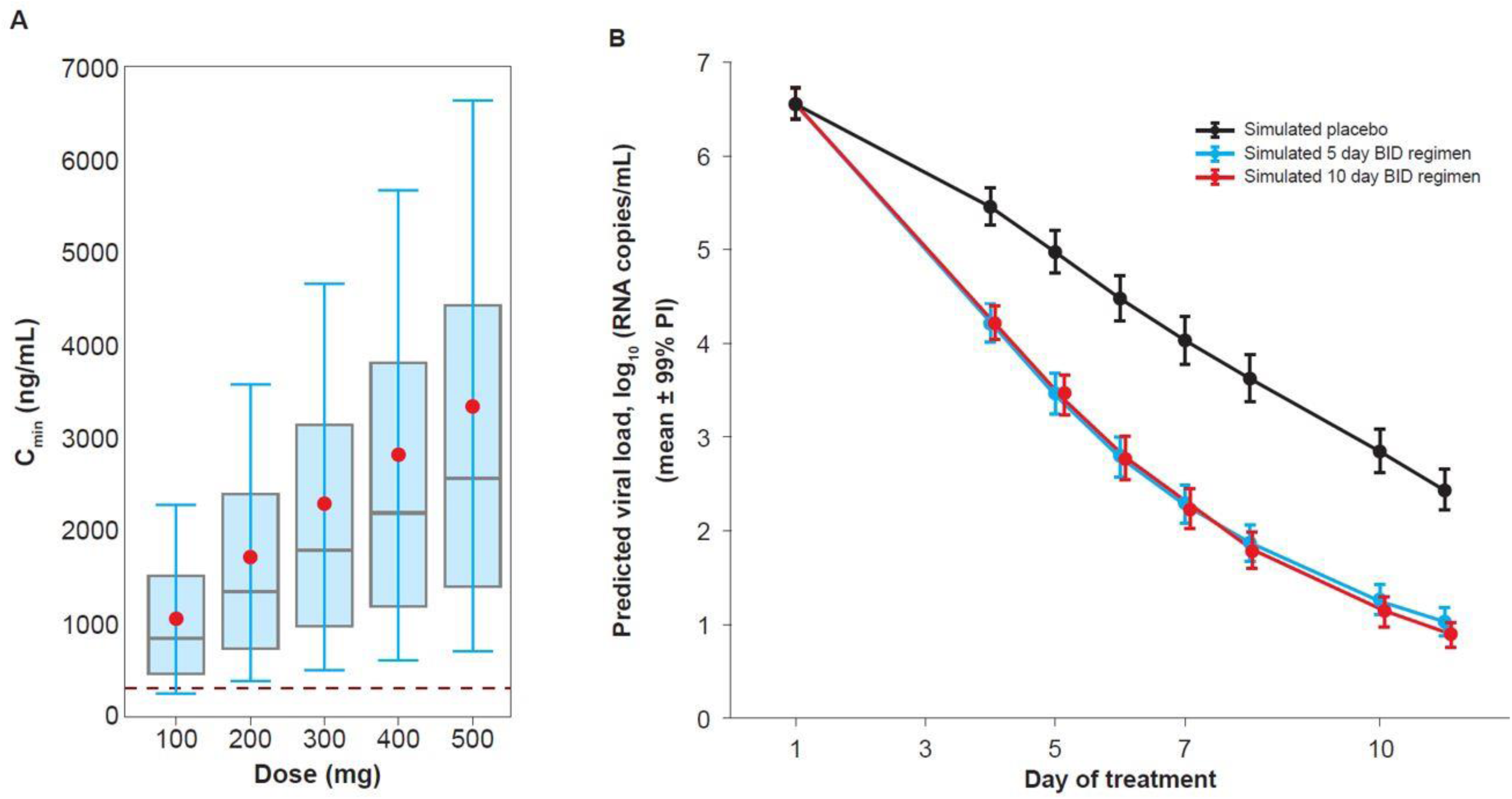
Distribution of simulated C_min_ at steady-state with different doses (BID) enhanced with ritonavir (A), and simulation of a virtual population (n=502) to predict viral load efficacy for nirmatrelvir/ritonavir 300/100 mg twice daily in symptomatic COVID-19 patients (B). In panel A, red dots indicate the means, grey lines indicate the medians, boxes are 25th and 75th percentiles, error bars show 10th and 90th percentiles, and the red dotted line is EC_90_ of 292 ng/mL. EC_90_= concentration at which 90% inhibition of viral replication is observed; PI= prediction interval; RNA=ribonucleic acid.

QSP model simulations of a virtual population treated with nirmatrelvir/ritonavir 300/100 mg BID (**Figure 3B**) predicted that 5 days of treatment is needed for robust viral reduction (based on clinical trials of monoclonal antibodies^21, 23^) in patients with symptomatic COVID-19, with no meaningful additional benefit with longer dosing.

## DISCUSSION

An urgent need exists for safe and effective COVID-19 therapeutics in community settings that can reduce viral load and transmission, improve time to clinical recovery, and prevent progression to adverse outcomes.^25–28^ Patient populations to benefit include those who are immunocompromised, unvaccinated, or are fully vaccinated but have waning immunity or breakthrough infections. At the time of this study, no orally administered antiviral had received EUA to treat non-hospitalized patients with COVID-19. On December 22, 2021, the US FDA issued an EUA for nirmatrelvir/ritonavir for the treatment of mild-to-moderate COVID-19 in patients 12 years and older who are at high risk of progression to severe disease, including hospitalization and death.^29^ The primary data supporting EUA for nirmatrelvir/ritonavir was from the phase 3, placebo-controlled EPIC-HR study, which showed nirmatrelvir/ritonavir to be 88% efficacious at reducing hospitalization or death, with no deaths recorded in the treatment group in patients who started treatment up to 5 days after symptom onset.^29, 30^ Molnupiravir, a nucleoside inhibitor, received EUA the following day.^31, 32^

Preclinical nirmatrelvir evaluations demonstrated potent antiviral efficacy, oral bioavailability, and a clean toxicity profile, making a compelling case for clinical development.^15^ In response to the urgent need for COVID-19 therapeutics, we used an innovative and seamless approach to generate safety and pharmacokinetic data to support rapid progression to phase 2/3 studies in patients with mild-to-moderate COVID-19, by designing this first-in-human study with multiple parts and flexibility. The interleaving design, rapid sample analysis, and implementation of automated data analysis allowed twice a week dose escalation, with the in-clinic phase of SAD and MAD completed within 16 days and 6 weeks, respectively, compared with approximately 6 months following a normal paradigm. The design allowed optional implementation of ritonavir as a pharmacokinetic enhancer based on emerging pharmacokinetic data. A cohort to study safety and tolerability at supratherapeutic exposure was added to reduce the need for ECG monitoring in phase 2/3 studies and demonstrate lack of pro-arrhythmic risk of nirmatrelvir.^33^ The study conduct and rapid adaption to emerging data together with near real-time data analysis and MIDD offered substantial decrease in the overall development and decision timelines.

Nirmatrelvir was safe and well tolerated in single-, multiple-ascending, and supratherapeutic dose cohorts. All treatment-emergent AEs were mild in severity. No severe AEs or SAEs were reported.

A pharmacokinetic enhancer (ritonavir)^34^ was used to maximize clinical concentrations to increase confidence in achieving clinical anti-viral efficacy through maintaining exposures multiple times above the in vitro EC_90_ and provide a potential barrier against resistance development. As indicated by the urinary excretion data, in the presence of ritonavir (limiting metabolism of nirmatrelvir), renal elimination was the major pathway of elimination. Coadministration resulted in an approximately 8-fold increase in nirmatrelvir concentrations. Simulations showed coadministration of nirmatrelvir 300 mg with ritonavir 100 mg maintained a free concentration at 12 hours post-dose (C_min_) above efficacious concentrations in >90% of potential trial participants from the very first dose, with median free C_min_ >6 times the in vitro EC_90_ at steady-state.^15^

The effect of food after oral administration of a nirmatrelvir suspension, when enhanced with ritonavir, was minimal. Given intended use of coadministration of nirmatrelvir with ritonavir, nirmatrelvir is being dosed without regard to food.

Population pharmacokinetic modeling and simulations enabled selection of the nirmatrelvir/ritonavir dose and dosing regimen for phase 2/3 (300/100 mg BID), predicted to achieve efficacious concentrations continuously in ≥90% of dosed participants. This target, which was successfully implemented for the first time for SARS-CoV-2 in the current study, was selected based on the literature, where exposure in multiples of EC_90_ for oral protease and non-nucleoside protease inhibitors correlates with clinical potency.^16, 17^ This was also supported by efficacy studies in pre-clinical species infected with SARS-CoV-2,^15^ and projected to translate to human antiviral and relative risk reduction benefits when applied in the QSP modeling. In fact, QSP modeling predicted that 5 days of dosing of oral nirmatrelvir/ritonavir 300/100 mg BID would be clinically efficacious, supporting this duration for phase 2/3 trials. Dosing for >5 days in symptomatic patients was predicted to have no additional benefit. The physiological rationale for this observation is that after 5 days of dosing (approximately 9 days after symptom onset and about 2 weeks after infection^21, 35, 36^) viral dynamics may be almost entirely driven by viral clearance rates, and not by viral replication.

Limitations of the study include that only a small number of participants were enrolled to provide an initial evaluation of safety and pharmacokinetics in healthy participants. Larger trials across various patient populations were needed to confirm nirmatrelvir/ritonavir safety and evaluate efficacy. Such large studies in patients with COVID-19 (Evaluation of Protease Inhibition for COVID-19 in High-Risk Patients [EPIC-HR], in Standard Risk Patients [EPIC-SR], and Post-Exposure Prophylaxis [EPIC-PEP]) along with clinical pharmacology studies (eg, in participants with renal or hepatic impairment), are either completed or ongoing. Study participants were also of limited ethnic diversity. However, a small number of Japanese participants were included, and no meaningful differences in safety and pharmacokinetic results were identified between Japanese and non-Japanese participants.

In conclusion, in response to the urgent need for COVID-19 treatments, our innovative, seamless, and efficient approach in this study enabled accelerated dose and regimen selection for use in late phase clinical studies within 6 weeks from the first dose in humans without compromising the quality of safety and pharmacokinetic assessment, while providing the appropriate confidence to initiate pivotal efficacy trials. Nirmatrelvir/ritonavir was safe and well tolerated in healthy participants. Ritonavir enhanced nirmatrelvir pharmacokinetics, hereby expected to achieve plasma concentrations and maintain C_min_ values in multiples of the in vitro EC_90_ when dosed BID at steady-state. These data enable high confidence in selection of a regimen of nirmatrelvir 300 mg in combination with ritonavir 100 mg administered BID over a 5-day period for phase 2/3 clinical trials in patients with COVID-19.

## Study Highlights

### What is the current knowledge on the topic?

Nirmatrelvir is a potent and specific inhibitor of M^pro^ enzyme activity and anti-viral activity across a diverse spectrum of coronaviruses. Administration in a murine SARS-CoV-2 model demonstrated dose-dependent reduction of pulmonary viral titers and reduced tissue pathology.

### What question did this study address?

This first-in-human study in healthy adults, which used an innovative and seamless operational and automated data analysis approach to evaluate safety, tolerability, and pharmacokinetics of nirmatrelvir and selected dose regimen, and duration for phase 2/3 studies.

### What does this study add to our knowledge?

The safety, tolerability and pharmacokinetics of nirmatrelvir in healthy adults and the dose selection is explained. This study shows an innovative approach to inform and expedite the development of nirmatrelvir.

### How might this change clinical pharmacology or translational science?

This provides an example of a seamless phase 1 program with MIDD-based decision making to support expedited start and conduct of phase 2/3 study with at least 10-fold reduction in timeline compared to industry standards.

## Data Sharing Statement

Upon request, and subject to review, Pfizer will provide the data that support the findings of this study. Subject to certain criteria, conditions and exceptions, Pfizer may also provide access to the related individual de-identified participant data. See https://www.pfizer.com/science/clinical-trials/trial-data-and-results for more information.

## Protocol Sharing Statement

The protocol for this study is available from the corresponding author upon request.

## Acknowledgments

The authors thank Tricia Newell, PhD, and Sheena Hunt, PhD, of ICON (Blue Bell, PA, USA), who wrote the first draft under direction from the authors, with funding from Pfizer Inc.

We would like to thank all the participants who volunteered for this study. We also thank all the study site personnel for their contributions to this study.

The authors would also like to acknowledge the significant number of Pfizer colleagues who have contributed to this first-in-human study. In particular, we acknowledge Scott White for his contribution to the study conduct, Joanne Salageanu for NCA analysis, Claudine Fredette and Geraldine Gigou for their contribution with study conduct, Karen Bartsch for study management, NH CRU and BR CRU staffs supporting the conduct of this study, CJ Musante for support of QSP modeling, Li Di for PBPK simulations, Amit Kalgutkar for the PDM contributions, Charlotte Allerton for strategic input, and Britton Boras for the assessment of efficacious concentrations.

## Author Contributions

R Singh, S Toussi, F Hackman, R Rao, R Allen, L Van Eyck, S Pawlak, A Anderson, M Binks, S Menon, G Nucci, and A Bergman designed the research. R Rao, R Allen, L Van Eyck, S Pawlak, E Kadar, F Clark, and H Shi performed the research. R Singh, S Toussi, F Hackman, P Chan, L Van Eyck, S Pawlak, A Anderson, G Nucci, and A Bergman analyzed the data. All authors were involved in writing the manuscript.

## SUPPLEMENTARY INFORMATION

### Inclusion and Exclusion Criteria

The study included 18‒60-year-old men and women who were determined to be healthy by medical evaluation. Participants were to have a body mass index (BMI) of 17.5‒30.5 kg/m^2^ and a total body weight of >50 kg. The Japanese cohort included Japanese participants who had 4 Japanese biologic grandparents who were born in Japan.

Key exclusion criteria included evidence or history of clinically significant hematological, renal, endocrine, pulmonary, gastrointestinal, cardiovascular, hepatic, psychiatric, neurological, or allergic disease; any condition possibly affecting drug absorption; or a positive test result for SARS-CoV-2 infection at the time of screening or on Day –1. Participants with a history of HIV, hepatitis B, or hepatitis C infection or a positive test at screening for HIV, hepatitis B surface antigen (HBsAg), hepatitis B core antibody (HBcAb), or hepatitis C antibody (HCVAb) were excluded, but a positive HBsAb test due to hepatitis B vaccination was allowed. Participants who had received a COVID-19 vaccine ≤7 days before screening or admission, or who were to be vaccinated with a COVID-19 vaccine at any time during the study confinement period were excluded. Participants with a screening supine blood pressure ≥140 mmHg (systolic) or ≥90 mmHg (diastolic) following ≥5 minutes of supine rest, a baseline 12-lead electrocardiogram with clinically relevant abnormalities, or abnormalities in clinical laboratory tests at screening (eg, aspartate aminotransferase, alanine aminotransferase, or total bilirubin ≥1.5 × upper limit of normal) were also excluded.

### Ethical Conduct of the Study

Conduct of the study was in accordance with ethical principles derived from the Declaration of Helsinki and in compliance with International Conference on Harmonisation Guidelines for Good Clinical Practice. All regulatory requirements were followed, including those affording greater protection to the safety of trial participants. The study protocols and any amendments and informed consent documents were approved by the institutional review board/ethics committee. Written informed consent was obtained from all participants before any study activity.

### Study Responsibilities

Pfizer was responsible for study design and conduct; data collection, analysis, and interpretation; and writing of this manuscript. Pfizer manufactured nirmatrelvir and placebo. All data were available to the authors, who vouch for its accuracy and completeness and to adherence of the studies to the protocols.

### Operational Conduct of Phase 1 Study

Initially, a 3-part study including SAD, MAD, and relative bioavailability and food effect cohort was designed, with the relative bioavailability and food effect cohort being optional. The SAD part was a randomized, double-blind, sponsor-open, placebo substitution design and included 2-interleaving cohorts for 4 periods with period 3 and 4 being optional. It also allowed flexibility to implement ritonavir and evaluate relative bioavailability and food effect, as feasible. The SAD part also included an exploratory objective to implement metabolite identification by a novel ^19^F-NMR method. The MAD part was a randomized, double-blind, placebo-controlled, sponsor-open design and included 5 cohorts including 1 Japanese and 2 non-Japanese cohorts being optional. The relative bioavailability and food effect part was an optional, randomized, 3-period, open-label cohort to evaluate relative bioavailability and food effect of a tablet formulation if it became available during the conduct of the trial.

Upon receipt of the US Food and Drug Administration approval on February 19, 2021, to conduct this study, the first participant in SAD was dosed on March 2, 2021 and the subsequent dosing occurred twice a week, every 3 to 4 days (every Tuesday and Friday) in Cohort 1 and 2 in an alternating manner. Pharmacokinetic samples collected up to 8 hours after dose on Day 1 were shipped on the same day to the bioanalytical lab. Sample analysis using a validated LC-MS/MS assay was performed and the pharmacokinetic concentrations became available the next day. Automated analysis of the cumulative pharmacokinetics and safety data (up to 48 hours post dose) was conducted and reviewed in the morning of Day 3 to enable a dose escalation decision and next day dosing of participants in the alternate cohort. Upon completion of the first 3 doses of nirmatrelvir alone (150 mg, 500 mg, and 1500 mg), it was evident that the exposure of nirmatrelvir did not increase with dose as expected, and pharmacokinetic enhancement with ritonavir was necessary to achieve C_min_ at multiples of EC_90_. Fit-for-purpose pharmacokinetic modeling (empirical and physiological-based pharmacokinetics) based on the available pharmacokinetic data from 3 doses and in vitro data were conducted to predict the exposure of nirmatrelvir in the presence of ritonavir. Supported by the modeling, a dose of nirmatrelvir (250 mg) when enhanced with ritonavir was selected to achieve approximately 3 times higher exposure than nirmatrelvir 1500 mg alone as the next escalation. Subsequently, the dose was escalated to nirmatrelvir 750 mg enhanced with ritonavir, and the safety and pharmacokinetics obtained supported initiation of the MAD part. Cohort-2, Period 3 was used to evaluate the effect of a high-fat, high-calorie meal on the pharmacokinetics of nirmatrelvir enhanced with ritonavir. Optional period 4 was not conducted.

Dosing in MAD (cohort 3) started on March 18, 2021, with dose escalation occurring approximately every week. Similar to SAD, the pharmacokinetic samples collected up to 8 hours post dose on Day 5 were shipped on the same day, analyzed, and the safety data up to Day 6 was reviewed before selecting a dose for the subsequent cohort. MAD cohorts ran in parallel allowing in-clinic assessment of safety and pharmacokinetics up to nirmatrelvir/ritonavir 500/100 mg BID by April 13, 2021. Data snapshots from SAD and MAD were taken on April 7 and 14, respectively, to enable start of the phase 2/3 study. Based on available preliminary pharmacokinetic data from SAD and MAD, a population pharmacokinetic model was developed, and simulations of different doses and dosing regimens were conducted to select the phase 2/3 dose. The simulated pharmacokinetic data from this population pharmacokinetic model also fed into the quantitative structural pharmacology model to support the dosing duration. Additionally, the preliminary data on relative bioavailability of a tablet formulation (250 mg) showed comparable exposure to the oral suspension (∼20% decrease). Based on all these data, a dose of nirmatrelvir/ritonavir 300/100 mg BID was selected on April 30, 2021, which was less than 2 months from the first clinical trial dose in the study.

The urinary pharmacokinetic data from MAD indicated the change in primary route of elimination from metabolism to renal when administered with ritonavir. Thus, a study assessing the impact of renal impairment (NCT04909853) was initiated.

An exploratory objective of evaluating the feasibility of ^19^F-NMR for metabolite identification provided us enough confidence in our ability to fulfil regulatory requirements using this novel method. Therefore, to expedite the drug development timeline, the protocol was amended on June 2, 2021, to include a metabolism and excretion cohort.

Upon feedback on June 16, 2021, from the FDA on the phase 2/3 protocol to include ECG monitoring citing insufficient exposure margins in our phase 1 program above the expected therapeutic C_max_, the phase 1 protocol was amended on June 24, 2021 to include a supratherapeutic exposure cohort to assess safety, tolerability and pharmacokinetics at exposures that were multiples of the expected C_max_ at the therapeutic dose. Given the less than proportional increase observed in SAD and MAD, increasing the dose was unlikely to achieve supratherapeutic exposure. Supported by modeling and simulations, a dose of 2250 mg was split into 3 doses of 750 mg administered at 0, 2 and 4 hours. In the supratherapeutic cohort, participants received the first dose on July 22, 2021 and the in-clinic phase was completed on August 1, 2021. With the rapid turnaround of the pharmacokinetic samples and implementation of automated concentration-QTc analysis, the data were submitted to the FDA on August 10, 2021, along with ECG data collected in the sentinel cohort of the phase 2/3 study, leading to the agency’s agreement to stop further ECG monitoring in phase 2/3 studies on August 26, 2021.

### Methods to Ensure Blinding

Investigators and participants were blinded to each participant’s assigned study intervention throughout the course of the study. To maintain this blind, an otherwise uninvolved third party (for example, pharmacist) was responsible for the preparation and dispensing of all study intervention according to the randomization schedule and assigned treatment for the individual participant. This third party instructed the participant to avoid discussing the taste, or packaging of the study intervention with the investigator.

Blood specimens were obtained from all participants for pharmacokinetic analysis to maintain the study blind at the investigator site. Only the investigator site staff and blinded study monitor, if assigned, were blinded to study treatment. A limited number of study team personnel were unblinded to participant treatments to permit real-time interpretation of the safety and pharmacokinetic data; and provided information necessary to potentially alter the dose escalation sequence. The blinded study monitor, if assigned, was blinded to treatment until all monitoring for the study was completed.

Specimens from participants randomized to placebo were not be routinely analyzed. To minimize the potential for bias, treatment randomization information was kept confidential by unblinded personnel and was not released to the blinded investigator or blinded investigator site personnel until the study database was locked or the investigator requested unblinding for safety reasons.

### Randomization

Independent pharmacists dispensed either nirmatrelvir or placebo according to a computer-generated randomization list which was provided to the study site by the sponsor. Participants were randomly assigned following simple randomization procedures using blocking.

---

**Figure S1.**
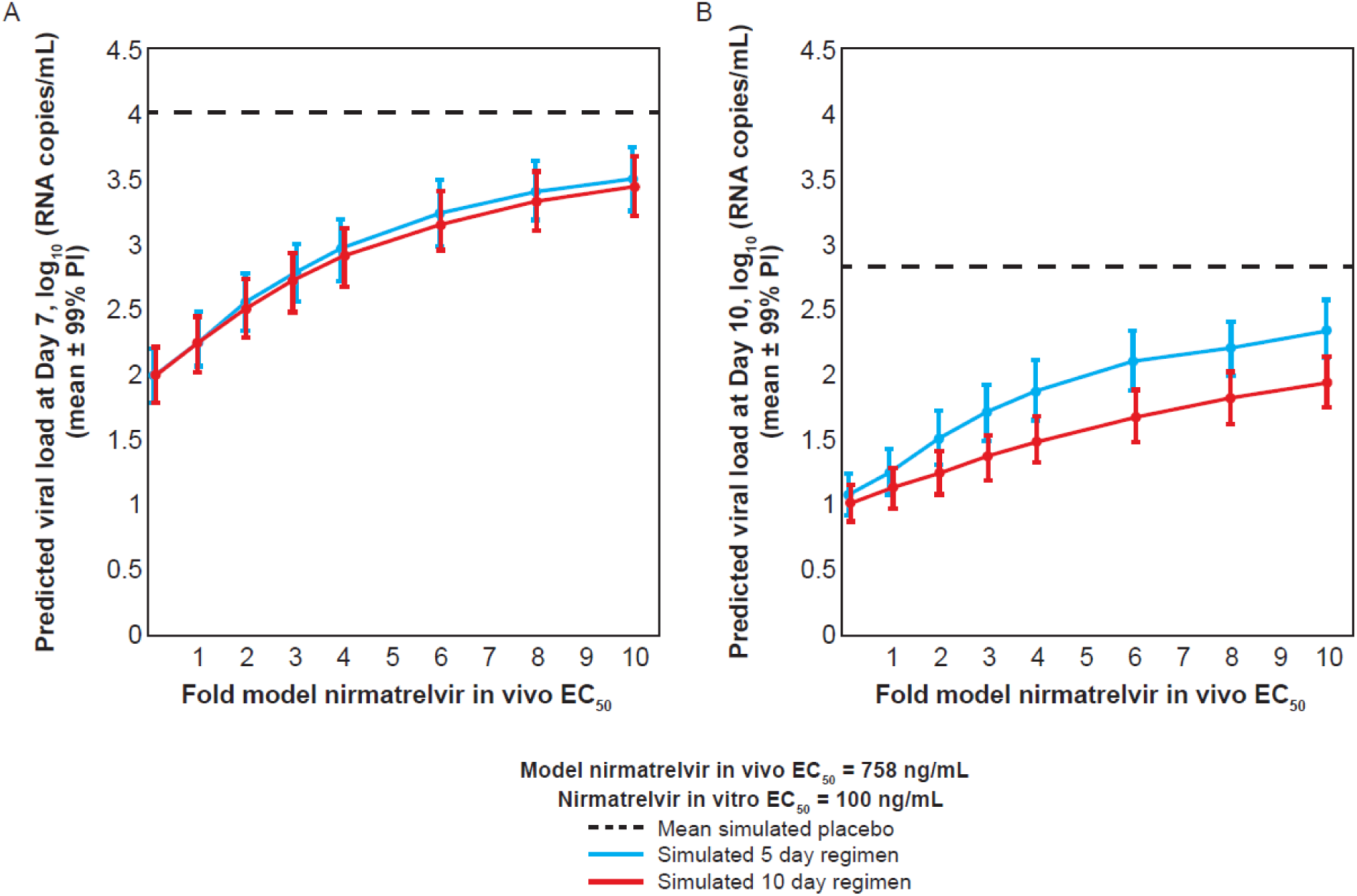
Simulation of a virtual population (n= 502) in the QSP model at day 7 (A) and day 10 (B) following 5 or 10 days of dosing with nirmatrelvir/ritonavir 300/100 mg twice daily. At day 10, a ∼0.4 log_10_ difference in viral load is predicted between the 5-day and 10-day dosing regimens for a greater than 3-fold shift in the estimated in vivo potency of nirmatrelvir. However, in this case, the predicted viral load for both the 5-day and 10-day dosing regimens is below the typical limits of quantification, and hence the difference is presumed to be not clinically meaningful or measurable. EC_50_= concentration at which 50% inhibition of viral replication is observed; ie, concentration required for 50% effect; PI = prediction interval; RNA=ribonucleic acid.

**Table S1.**
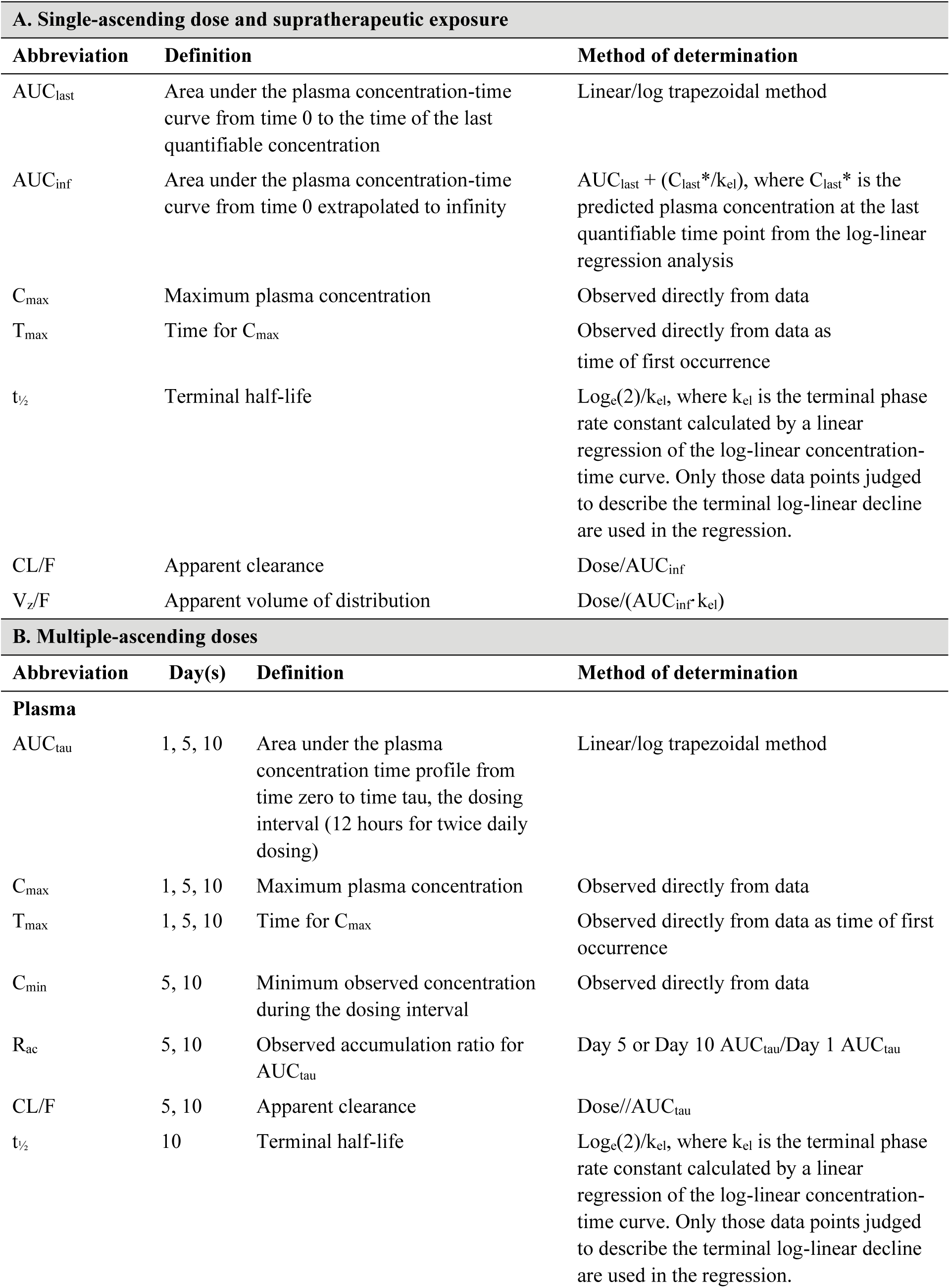
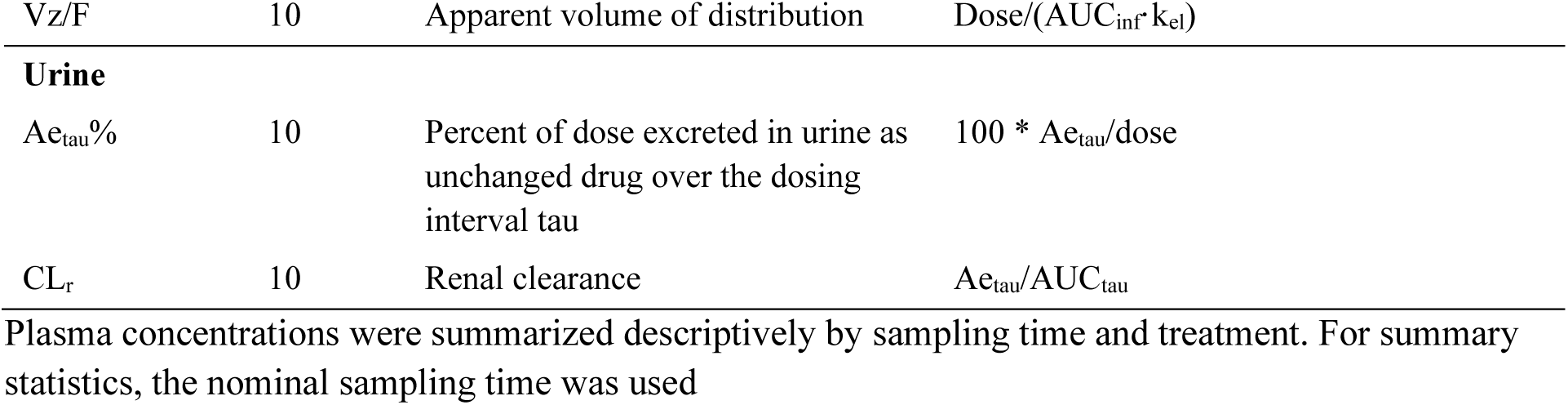
Derivation and definitions of the pharmacokinetic parameters (A) single-ascending dose, supratherapeutic exposure, and food effect and bioavailability, and (B) multiple-ascending doses.

**Table S2.** Study populations.

| **Population** | **Description** |
| --- | --- |
| Evaluable | All participants who were randomly assigned to study intervention and who took $\geq 1$ dose of study intervention for the given part of the study |
| Safety | All participants who were randomly assigned to study intervention and who took $\geq 1$ dose of study intervention |
| Pharmacokinetic parameter set | All participants randomly assigned to study intervention and who took $\geq 1$ dose of study intervention and in whom $\geq 1$ of the PK parameters of interest are reported for the given part of the study |

**Table S3.**
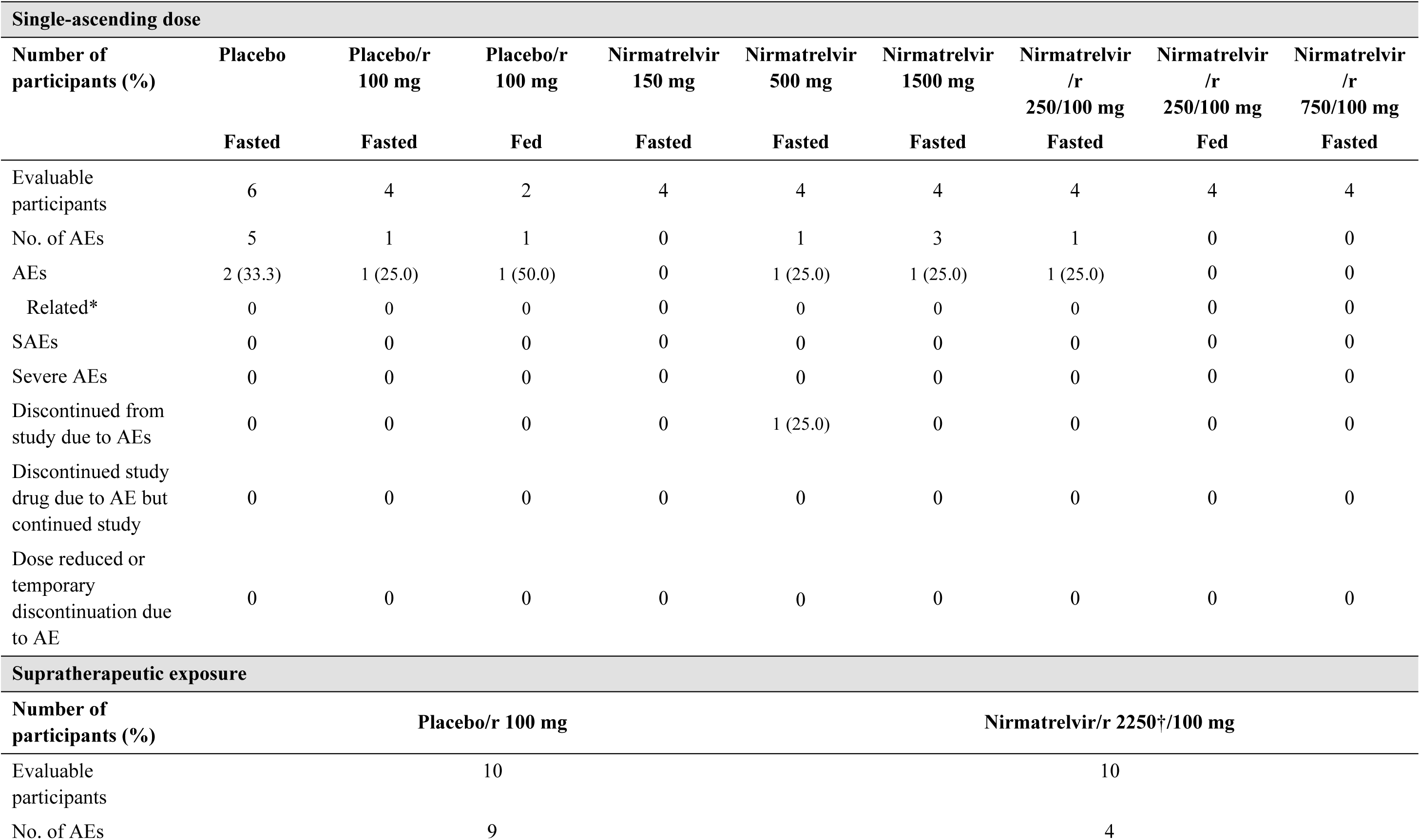
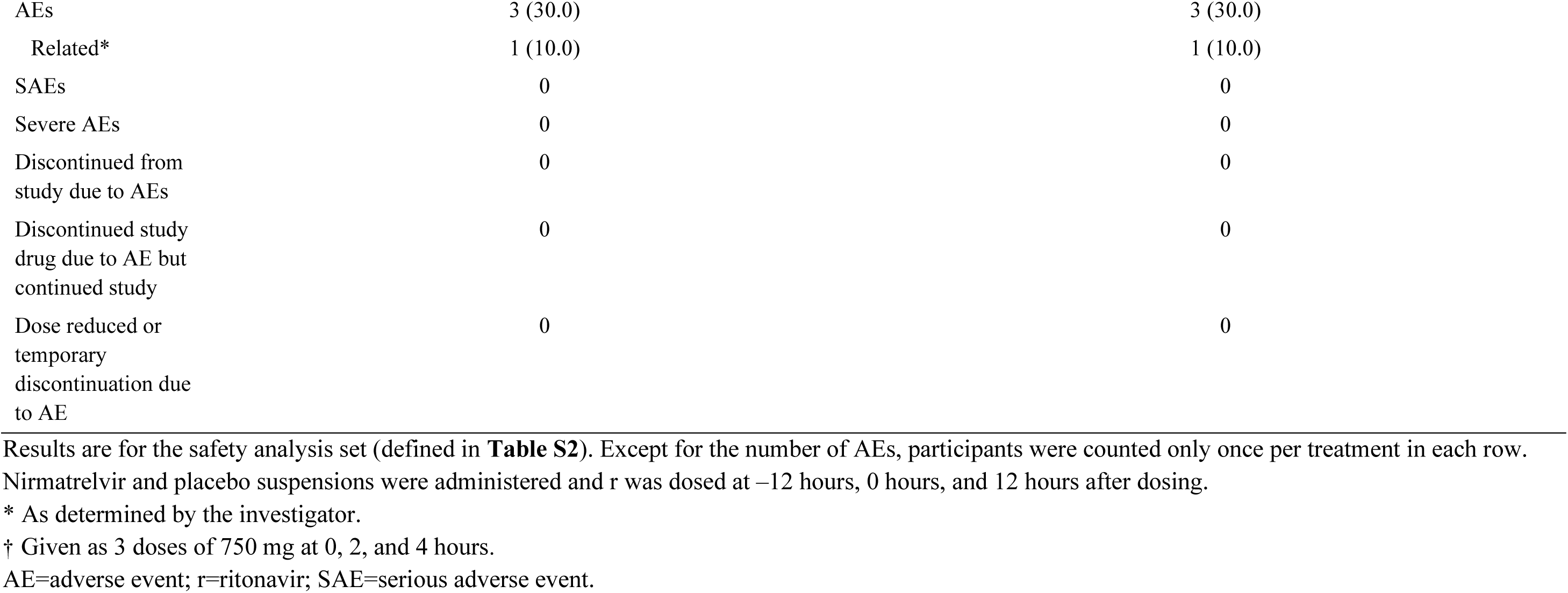
Treatment-emergent adverse events.

**Table S4.** Treatment-emergent adverse events in the multiple-ascending dose cohort.

| **Overall** |  |  |  |  |  |  |
| --- | --- | --- | --- | --- | --- | --- |
| **Number of participants (%)** | **Placebo/r**<br>**100 mg**<br>**Fasted** | **Placebo/r**<br>**100 mg**<br>**Fasted, Japanese** | **Nirmatrelvir/r**<br>**75/100 mg**<br>**Fasted** | **Nirmatrelvir/r**<br>**250/100 mg**<br>**Fasted** | **Nirmatrelvir/r**<br>**250/100 mg**<br>**Fasted, Japanese** | **Nirmatrelvir/r**<br>**500/100 mg**<br>**Fasted** |
| Evaluable participants | 8 | 2 | 4 | 4 | 4 | 7 |
| No. of AEs | 5 | 4 | 7 | 12 | 8 | 11 |
| AEs | 4 (50.0) | 2 (100.0) | 3 (75.0) | 3 (75.0) | 4 (100.0) | 4 (57.1) |
| Related* | 1 (12.5) | 1 (50.0) | 0 | 2 (50.0) | 2 (50.0) | 2 (28.6) |
| SAEs | 0 | 0 | 0 | 0 | 0 | 0 |
| Severe AEs | 0 | 0 | 0 | 0 | 0 | 0 |
| Discontinued from study due to AEs | 0 | 0 | 0 | 0 | 0 | 0 |
| Discontinued study drug due to AE but continued study | 0 | 0 | 0 | 0 | 0 | 0 |
| Dose reduced or temporary discontinuation due to AE | 0 | 0 | 0 | 0 | 0 | 0 |
| **Treatment-related adverse events\* by preferred term** |  |  |  |  |  |  |
| Blood TSH increased |  |  |  |  |  |  |
| Mild | 1 (12.5) | 1 (50.0) | 0 | 1 (25.0) | 1 (25.0) | 1 (14.3) |
| Moderate | 0 | 0 | 0 | 0 | 0 | 0 |
| Severe | 0 | 0 | 0 | 0 | 0 | 0 |
| Dysgeusia |  |  |  |  |  |  |
| Mild | 0 | 0 | 0 | 1 (25.0) | 1 (25.0) | 1 (14.3) |
| Moderate | 0 | 0 | 0 | 0 | 0 | 0 |
| Severe | 0 | 0 | 0 | 0 | 0 | 0 |

**Table S5.** Single-ascending dose assessment of plasma nirmatrelvir pharmacokinetic parameters: descriptive summary (A) and statistical summary of food effect (B)

| **A. Descriptive summary** |  |  |  |  |  |  |
| --- | --- | --- | --- | --- | --- | --- |
| **Parameter (unit)\*** | **Nirmatrelvir 150 mg**<br>**Fasted (N=4)** | **Nirmatrelvir 500 mg**<br>**Fasted (N=4)** | **Nirmatrelvir 1500 mg**<br>**Fasted (N=4)** | **Nirmatrelvir/r**<br>**250/100 mg**<br>**Fasted (N=4)** | **Nirmatrelvir/r**<br>**250/100 mg**<br>**Fed (N=4)** | **Nirmatrelvir/r**<br>**750/100 mg**<br>**Fasted (N=4)** |
| N1, N2* | 4, 3 | 4, 2 | 4, 0 | 4, 4 | 4, 4 | 4, 4 |
| AUC<sub>last</sub>, ng·h/mL | 2125 (34) | 3753 (29) | 10870 (47) | 27,600 (13) | 28,020 (16) | 64,230 (39) |
| CL/F, L/h | 66.83 (43) | 91.2, 91.8† | NR | 8.865 (14) | 8.735 (17) | 11.22 (45) |
| V<sub>z</sub>/F, L | 190.6 (36) | 2440, 3390† | NR | 87.98 (28) | 73.48 (47) | 181.9 (35) |
| **B. Statistical summary – food effect** |  |  |  |  |  |  |
| **Parameter (unit)\*** | **Nirmatrelvir/r**<br>**250/100 mg fed (test)** | **Nirmatrelvir/r**<br>**250/100 mg fasted (test)** | **Ratio (%) (test/reference)**<br>**of adjusted geometric means\*** |  | **90% CI (%) for ratio†** |  |
| AUC<sub>last</sub>, ng·h/mL/mg | 28,020 | 27,600 | 101.5 |  | (90.18, 114.31) |  |

The statistical summary of food effect used mixed effect models with treatment as a fixed effect and participant as a random effect, which were applied to the natural log transformation of AUC<sub>last</sub> using participants with data from both periods only.
\* N1 is the number of participants contributing to the summary statistics and N2 is the number of participants where CL/F and V<sub>z</sub>/F were determined.
† Ratios (and 90% CIs) are expressed as percentages.
The parameters are defined in **Table S1**.
CI=confidence interval; NR=not reported; r=ritonavir.

**Table S6.**
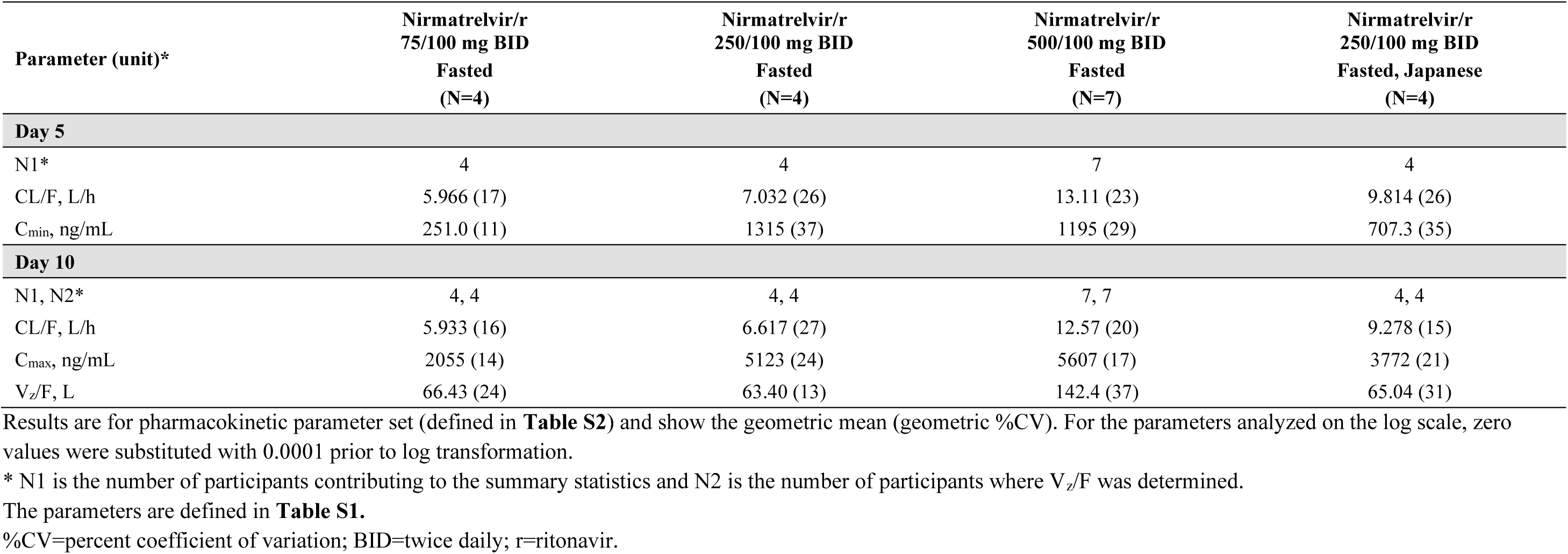
Multiple-ascending dose assessment: descriptive summary of plasma nirmatrelvir pharmacokinetic parameters.

**Table S7.** Descriptive summary of plasma nirmatrelvir pharmacokinetic parameters for supratherapeutic effect.

| Parameter (unit) | Nirmatrelvir/r 2250*/100 mg<br>(N=10) |
| --- | --- |
| N1, N2† | 10, 10 |
| AUC<sub>last</sub>, ng·h/mL | 188,200 (35) |
| CL/F, L/h | 3.970 (35) |
| t<sub>1/2</sub>, h | 7.450 ± 2.9357 |
| V<sub>z</sub>/F, L | 40.06 (55) |

\* Given as 3 doses of 750 mg at 0, 2, and 4 hours.
† N1 is the number of participants contributing to the summary statistics and N2 is the number of participants where t<sub>1/2</sub>, CL/F and V<sub>z</sub>/F were determined.
The parameters are defined in **Table S1**.

**Table S8.** Predicted C_min_ in ng/mL and percentage of simulated subjects achieving C_min_ ≥EC_90_ of 292 ng/mL (variability in CL assumed to be 60%)

| Dose (mg)<br>plus ritonavir | Time point | Median | 10th Percentile | 90th Percentile | Participants achieving<br>$C_{min} \geq EC_{90}$ (%) |
| --- | --- | --- | --- | --- | --- |
| 100 | First dose | 458 | 141 | 1018 | 71.5 |
|  | Steady state | 852 | 238 | 2276 | 85.3 |
| 200 | First dose | 743 | 228 | 1608 | 85.0 |
|  | Steady state | 1361 | 383 | 3575 | 93.4 |
| 300 | First dose | 987 | 307 | 2124 | 90.7 |
|  | Steady state | 1800 | 498 | 4670 | 95.7 |
| 400 | First dose | 1209 | 378 | 2565 | 94.0 |
|  | Steady state | 2197 | 605 | 5679 | 97.4 |
| 500 | First dose | 1417 | 449 | 2979 | 95.5 |
|  | Steady state | 2563 | 704 | 6640 | 97.8 |

CL=clearance; $C_{min}$ =minimum concentration during the dosing interval (12 hours); $EC_{90}$ =concentration at which 90% inhibition of viral replication is observed; ie, concentration required for 90% effect.

